## Supplementary Materials for "Genetic surveillance in the Greater Mekong Subregion and South Asia to support malaria control and elimination"

### SUPPLEMENTARY TABLES

#### Supplementary Table 1 - Geographical breakdown by year of samples processed by GenRe-Mekong

| Region | Code | Year of Collection | | | | | | | | Total |
|---|---|---|---|---|---|---|---|---|---|---|
| | | 2012 | 2013 | 2014 | 2015 | 2016 | 2017 | 2018 | 2019 | |
| **Bangladesh** | BD | | | | 949 | 1210 | 24 | | | 2183 |
| **DR Congo** | CD | | | | 41 | 191 | | | | 232 |
| **India** | IN | | | | | 83 | 208 | 12 | | 303 |
| **Cambodia** | KH | 116 | 178 | 257 | 152 | 354 | 190 | 15 | | 1262 |
| **Lao PDR** | LA | | | | 11 | 4 | 680 | 875 | | 1570 |
| **Myanmar** | MM | | | | 112 | 791 | 457 | | | 1360 |
| **Thailand** | TH | | 9 | 7 | 82 | 20 | 51 | 77 | | 246 |
| **Vietnam** | VN | | | | 36 | 276 | 888 | 1126 | 141 | 2467 |
| | **Total** | 116 | 187 | 264 | 1383 | 2929 | 2498 | 2105 | 141 | **9623** |

**Supplementary Table 2 - Counts of processed samples, by province/state/division of origin.**

| Country | Province, State or Division | # samples processed |
|---|---|---|
| **India** | Odisha | 116 |
| | West Bengal | 103 |
| | Tripura | 84 |
| **Bangladesh** | Chittagong | 2183 |
| **Myanmar** | Rakhine | 56 |
| | Bago | 109 |
| | Mandalay | 125 |
| | Kayin | 1070 |
| **Thailand** | Tak | 30 |
| | Sisakhet | 172 |
| | Ubon Ratchathani | 44 |
| **Cambodia** | Pailin | 133 |
| | Battambang | 69 |
| | Pursat | 473 |
| | Preah Vihear | 142 |
| | Stueng Traeng | 78 |
| | Ratanakiri | 367 |
| **Lao PDR** | Champasak | 271 |
| | Attapeu | 315 |
| | Sekong | 47 |
| | Salavan | 210 |
| | Savannakhet | 727 |
| **Vietnam** | Binh Phuoc | 967 |
| | Dak Nong | 141 |
| | Dak Lak | 288 |
| | Gia Lai | 882 |
| | Khanh Hoa | 78 |
| | Ninh Thuan | 62 |
| | Quang Tri | 49 |
| **DR Congo** | Kinshasa | 232 |
| | **Grand Total** | **9623** |

**Supplementary Table 3 - Number of samples carrying mutations in the resistance domains of *kelch13* by province/state/division.** Mutations found only in single samples are grouped together in the "singleton" column. For clarity, heterozygous samples (i.e. samples where both mutant and wild-type parasites were detected) are excluded.

| Country | Province, State or Division | None | C580Y | P441L | F446I | R561H | G449A | M562I | M476I | Y493H | R539T | P553L | C469F | G533S | G538V | P574L | A578S | Singleton | Total |
|---|---|---|---|---|---|---|---|---|---|---|---|---|---|---|---|---|---|---|---|
| **India** | **Odisha** | 79 | | | | | | | | | | | | | | | | | **79** |
| | **West Bengal** | 79 | | | | | | | | | | | | | | | 1 | | **80** |
| | **Tripura** | 66 | | | | | | | | | | | | | | | | | **66** |
| **Bangladesh** | **Chittagong** | 1523 | | | | | | | | | | | | | | | 5 | 1 | **1529** |
| **Myanmar** | **Rakhine** | 43 | | | | | | | | | | | | | | | | | **43** |
| | **Bago** | 33 | | | | | | | | | | | | | | | | | **33** |
| | **Mandalay** | 41 | | | 8 | 8 | | | | | | | | | | | | 1 | **58** |
| | **Kayin** | 318 | 18 | 110 | 75 | 53 | 55 | 38 | 36 | | | | 5 | 5 | 8 | | | 4 | **725** |
| **Thailand** | **Tak** | 9 | 13 | | | | | | | | | | | 3 | | | | 2 | **27** |
| | **Sisakhet** | | 80 | | | | | | | | | | | | | | | | **80** |
| | **Ubon Ratchathani** | 3 | 14 | | | | | | | | 2 | | | | | | | | **19** |
| **Cambodia** | **Pailin** | 5 | 62 | | | | | | | | | | | | | | | | **67** |
| | **Battambang** | | 31 | | | | | | | | | | | | | | | | **31** |
| | **Pursat** | 38 | 262 | | | | | | | 18 | 1 | | | | | | | 1 | **320** |
| | **Preah Vihear** | 38 | 68 | | | | | | | 2 | 1 | | | | | | | | **109** |
| | **Steung Treng** | 5 | 66 | | | | | | | | 1 | | | | | | | | **72** |
| | **Ratanakiri** | 170 | 149 | | | | | | | 2 | 1 | 3 | | | | | | | **325** |
| **Laos** | **Champasak** | 78 | 143 | | | | | | | | 2 | | | | | | | | **223** |
| | **Attapeu** | 142 | 112 | | | | | | | | 10 | | | | | | | | **264** |
| | **Sekong** | 21 | 2 | | | | | | | 3 | | | | | | | | | **26** |
| | **Salavan** | 153 | 28 | | | | | | | | | | | | | 2 | | | **183** |
| | **Savannakhet** | 558 | 43 | | | | | | | 10 | | | | | | 5 | | | **616** |
| **Vietnam** | **Binh Phuoc** | 51 | 579 | | | | | | | | | | | | | | | | **630** |
| | **Dak Nong** | 7 | 111 | | | | | | | | | | | | | | | | **118** |
| | **Dak Lak** | 9 | 214 | | | | | | | | | | | | | | | | **223** |
| | **Gia Lai** | 120 | 611 | | | | | | | | 1 | 1 | 4 | | | | | | **737** |
| | **Khanh Hoa** | 53 | 2 | | | | | | | | | 13 | | | | | | | **68** |
| | **Ninh Thuan** | 21 | 3 | | | | | | | | | | | | | | | | **24** |
| | **Quang Tri** | 32 | 6 | | | | | | | | | | | | | | | | **38** |
| **Congo PDR** | **Kinshasa** | 116 | | | | | | | | | | | | | | | 1 | | **117** |
| | **Total** | **3811** | **2617** | **110** | **83** | **61** | **55** | **38** | **36** | **35** | **19** | **17** | **9** | **8** | **8** | **7** | **7** | **9** | **6930** |

**Supplementary Table 4 Drug Resistance Frequencies in Vietnam.**
The table shows estimates of the proportion of parasites predicted to be *resistant* to different drugs in 20 districts of 7 provinces of Vietnam where ≥10 samples were collected.

| Province | District | Sample Count | ART-R | PPQ-R | DHA-PPQ -R | CQ-R | PYR-R | SD-R |
|---|---|---|---|---|---|---|---|---|
| **Binh Phuoc** | Bu Dop | 17 | 93% | 100% | 92% | 100% | 100% | 100% |
| | Bu Dang | 18 | 93% | 100% | 90% | 100% | 100% | 100% |
| | Bu Gia Map | 354 | 94% | 91% | 84% | 100% | 100% | 100% |
| **Dak Lak** | Ea Kar | 29 | 100% | 100% | 100% | 100% | 100% | 100% |
| | Ea Hleo | 33 | 93% | 93% | 84% | 100% | 100% | 100% |
| | Ea Sup | 33 | 96% | 89% | 81% | 100% | 100% | 100% |
| | Buon Don | 53 | 94% | 86% | 78% | 100% | 100% | 100% |
| | Dak Lak | 111 | 97% | 88% | 85% | 99% | 100% | 99% |
| **Dak Nong** | Cu Jut | 60 | 93% | 94% | 88% | 100% | 100% | 95% |
| | Tuy Duc | 64 | 95% | 91% | 87% | 100% | 100% | 100% |
| **Gia Lai** | Kong Chro | 10 | 40% | 50% | 33% | 100% | 100% | 70% |
| | Ia Grai | 47 | 87% | 84% | 83% | 100% | 100% | 98% |
| | Ia Pa | 87 | 26% | 33% | 20% | 99% | 100% | 99% |
| | Gia Lai | 90 | 77% | 70% | 64% | 93% | 99% | 92% |
| | Duc Co | 135 | 84% | 89% | 77% | 100% | 100% | 100% |
| | Krong Pa | 437 | 97% | 92% | 90% | 100% | 100% | 99% |
| **Khanh Hoa** | Khanh Vinh | 67 | 23% | 5% | 2% | 95% | 100% | 97% |
| **Ninh Thuan** | Bac Ai | 43 | 19% | 25% | 0% | 8% | 100% | 100% |
| **Quang Tri** | Dakrong | 11 | 0% | 0% | 0% | 73% | 91% | 30% |
| | Dong Ha | 30 | 26% | 11% | 0% | 70% | 71% | 68% |
| Grand Total | | 1,729 | 84% | 82% | 73% | 97% | 99% | 98% |

### SUPPLEMENTARY FIGURES

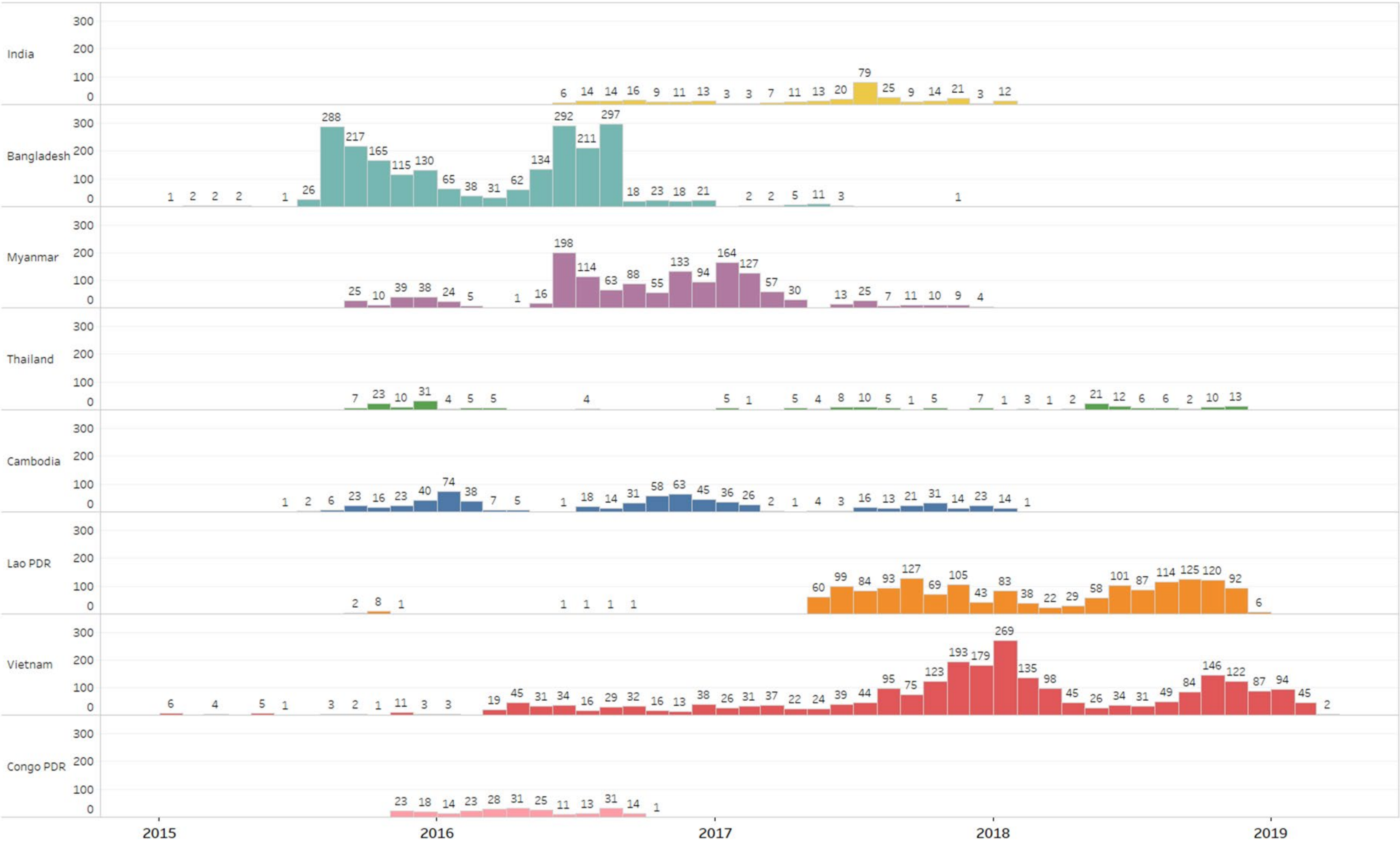

**Supplementary Figure 1 – Number of samples collected prospectively by month in each country.**

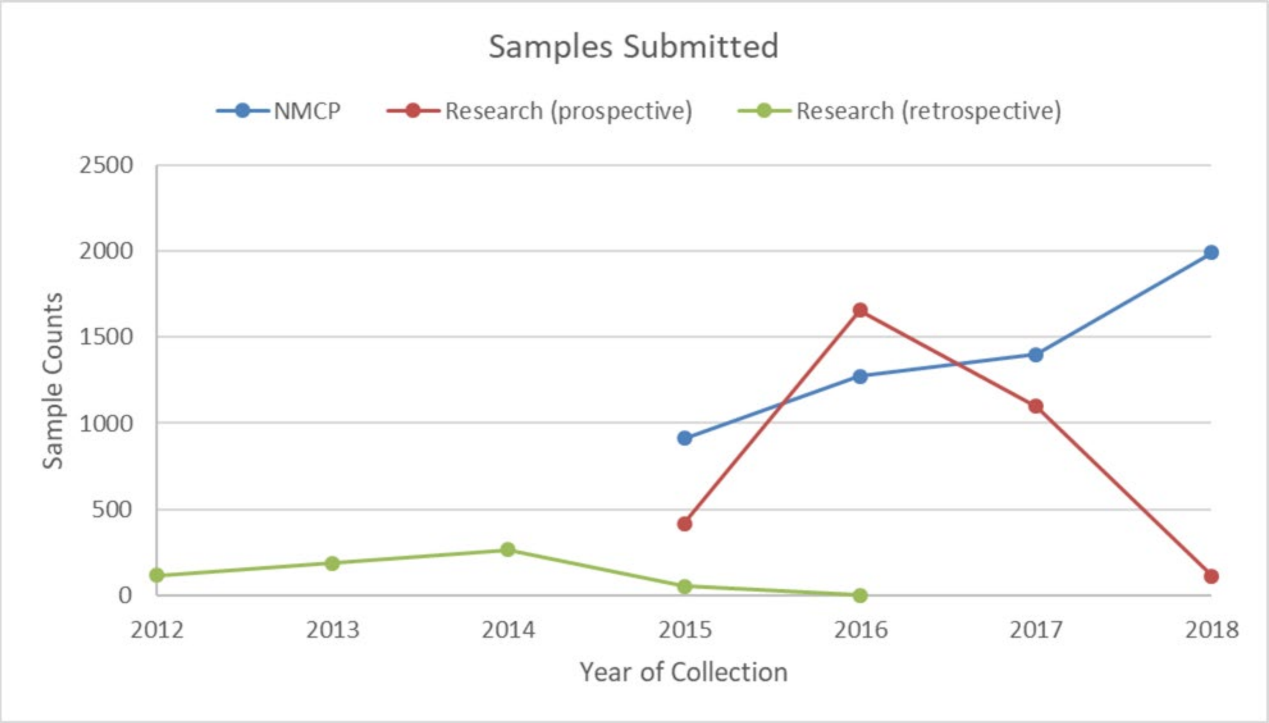

**Supplementary Figure 2 - Trends in sample collections over time.**

Numbers of samples collected prospectively each year by surveillance projects (blue) and research studies (orange) are compared. Sample counts submitted retrospectively by research projects (green) are also shown.

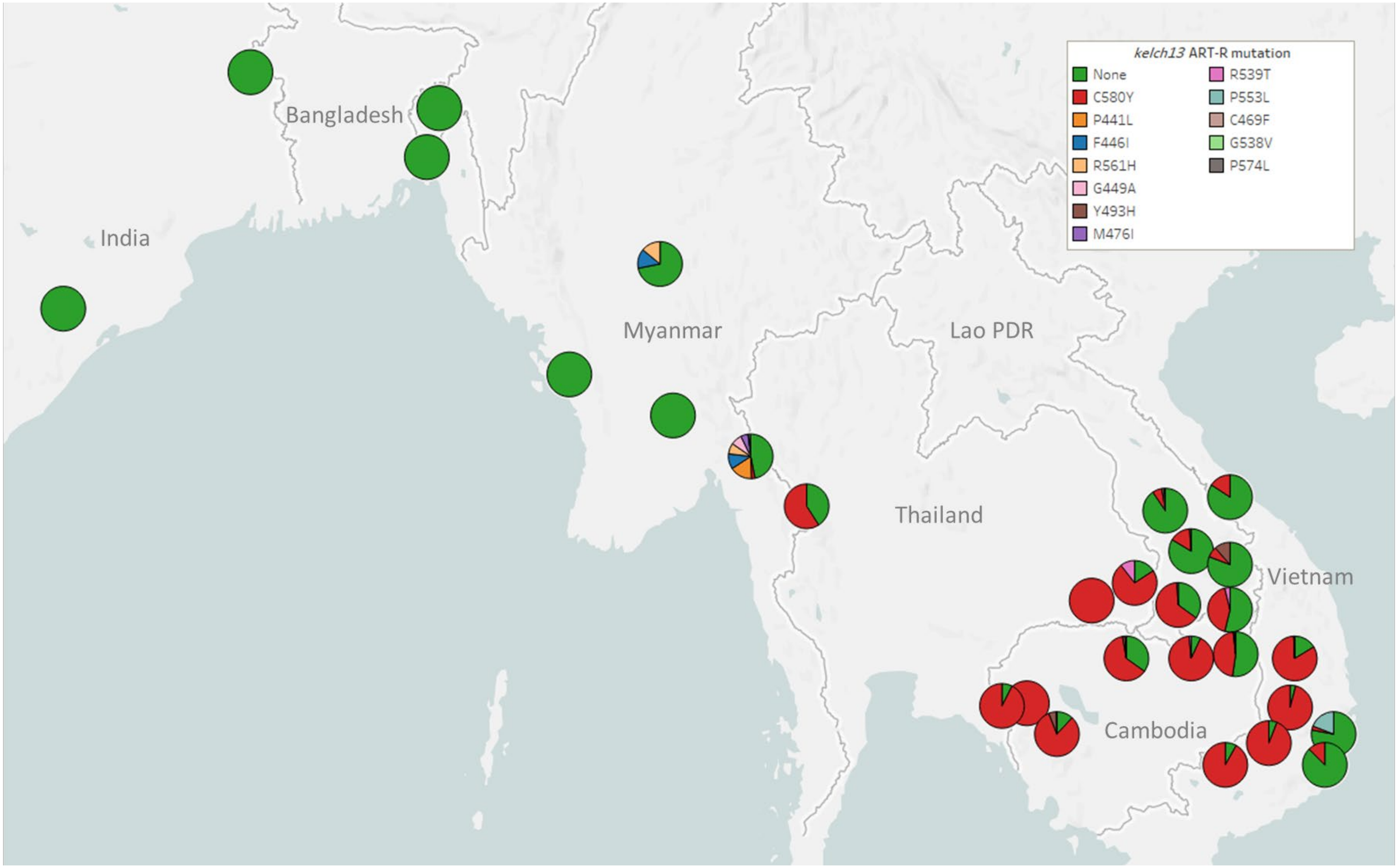

**Supplementary Figure 3 - kelch13 allele diversity in Asian countries.**

We show a pie chart for each province/state/division surveyed, indicating the relative proportion of different nonsynonymous mutations found in the resistance domains of *kelch13*. A total of 6,758 samples were included in this analysis, after excluding samples where the *kelch13* genotype could not be called, and those with *undetermined* ART-R phenotype prediction. For display clarity, mutations that we only found in singleton samples are also excluded (n=18).

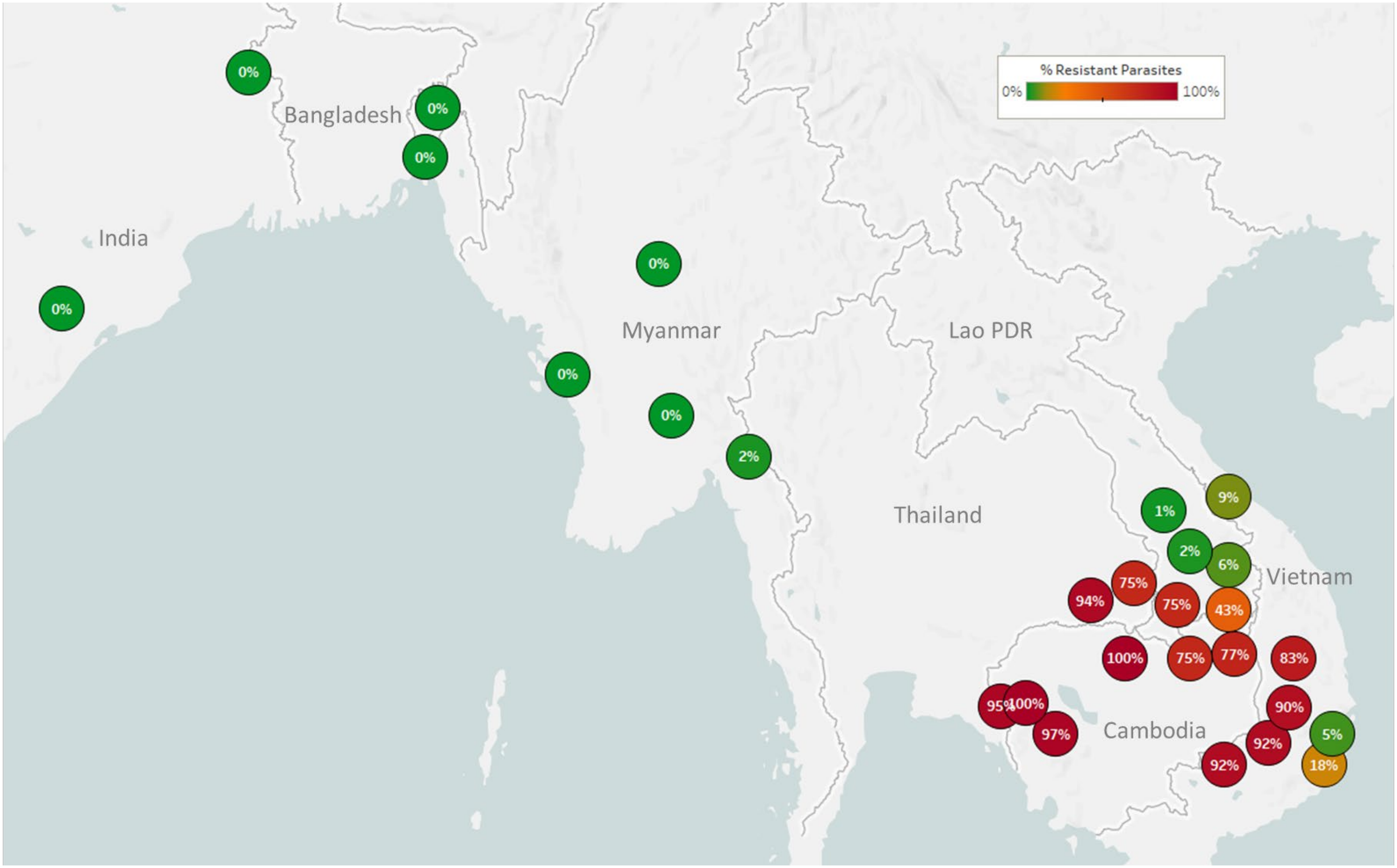

**Supplementary Figure 4 - Map of Piperaquine Resistance (PPQ-R) in Asian countries.**

Marker text and colour indicate the proportion of sample classified as *resistant* in each province/state/division surveyed. A total of 3,552 samples were included in this analysis, after excluding samples where *plasmepsin* 2/3 copy number could not be determined. The results are summarized in Table 3.

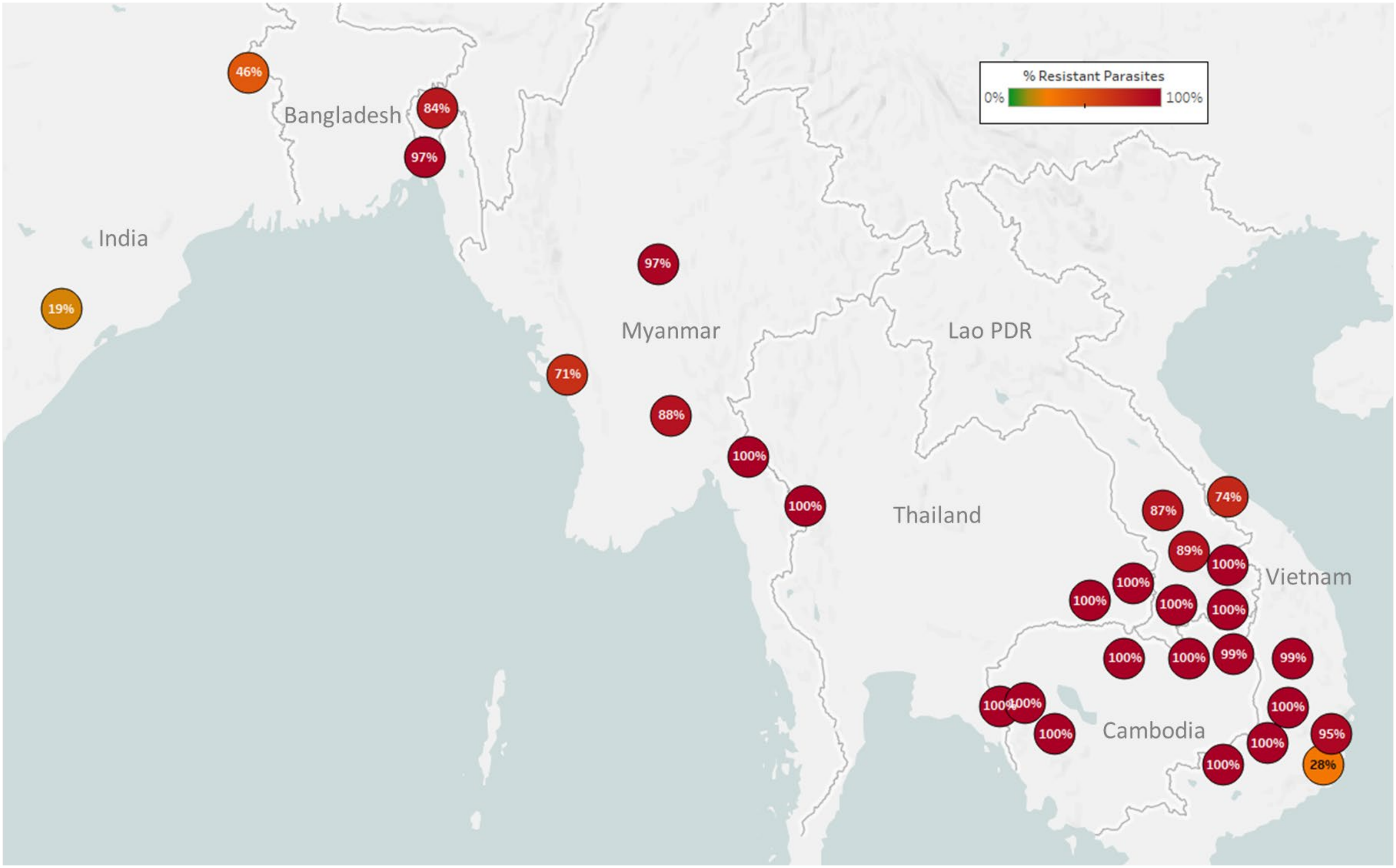

**Supplementary Figure 5 - Map of Chloroquine Resistance (CQ-R) in Asian countries.**

Marker text and colour indicate the proportion of sample classified as *resistant* in each province/state/division surveyed. A total of 6,458 samples were included in this analysis, after excluding samples where the *crt* core haplotype could not predict a phenotype. The results are summarized in Table 3.

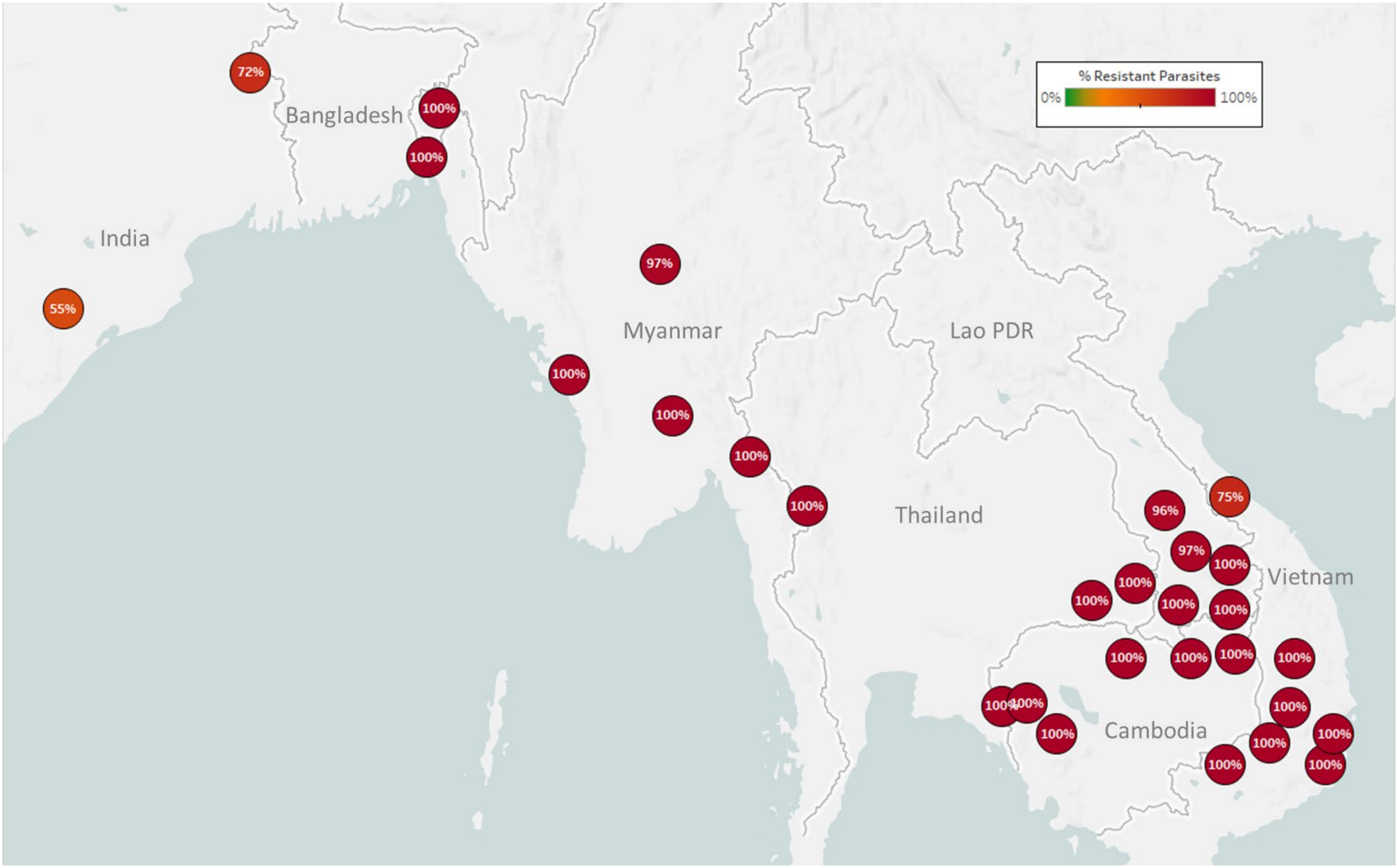

**Supplementary Figure 6 - Map of Pyrimethamine Resistance (PYR-R) in Asian countries.**

Marker text and colour indicate the proportion of sample classified as *resistant* in each province/state/division surveyed. A total of 7,208 samples were included in this analysis, after excluding samples where the *dhfr* core haplotype could not predict a phenotype. The results are summarized in Table 3.

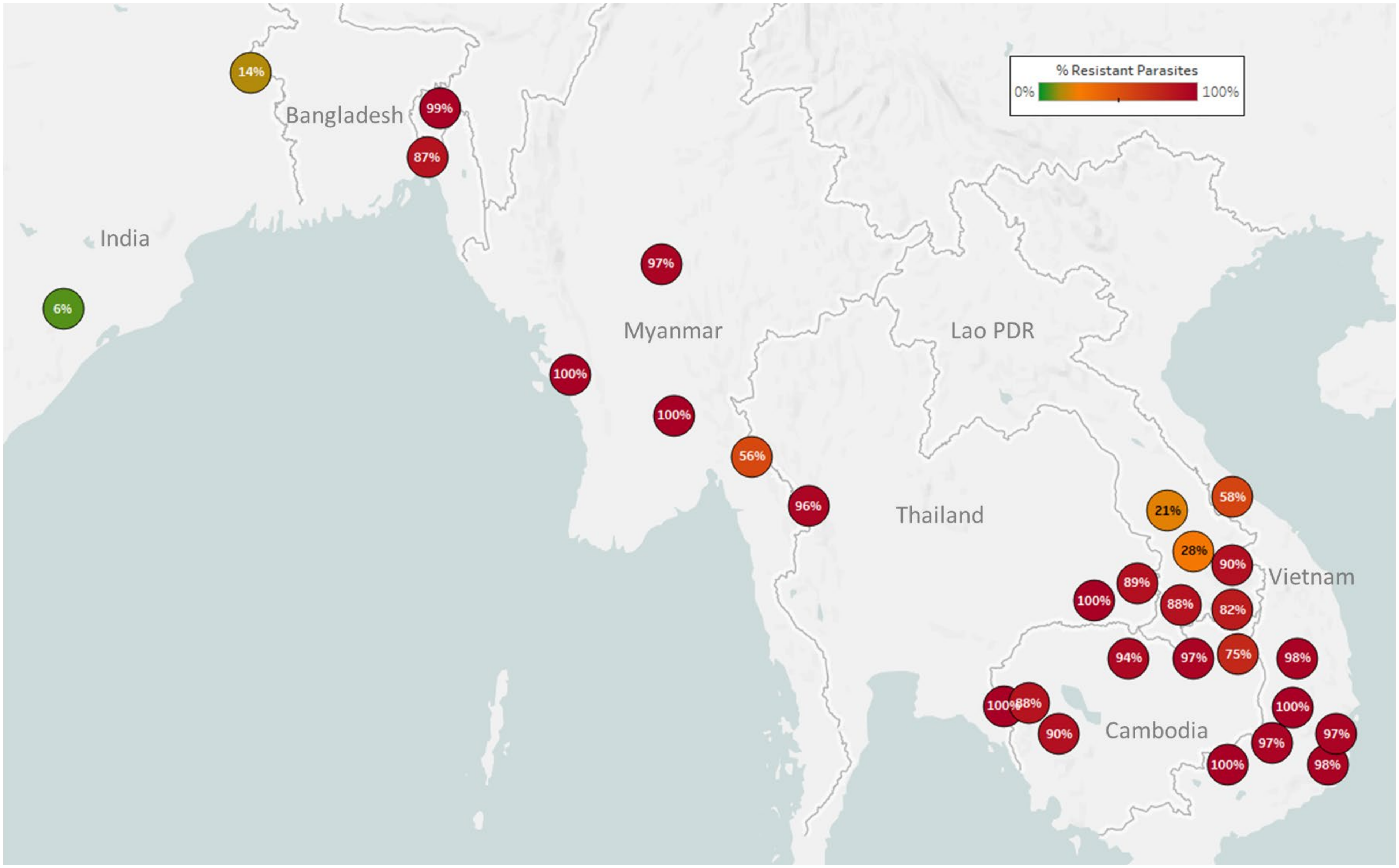

**Supplementary Figure 7 - Map of Sulfadoxine Resistance (SD-R) in Asian countries.**

Marker text and colour indicate the proportion of sample classified as *resistant* in each province/state/division surveyed. A total of 7,095 samples were included in this analysis, after excluding samples where the *dhps* core haplotype could not predict a phenotype. The results are summarized in Table 5.

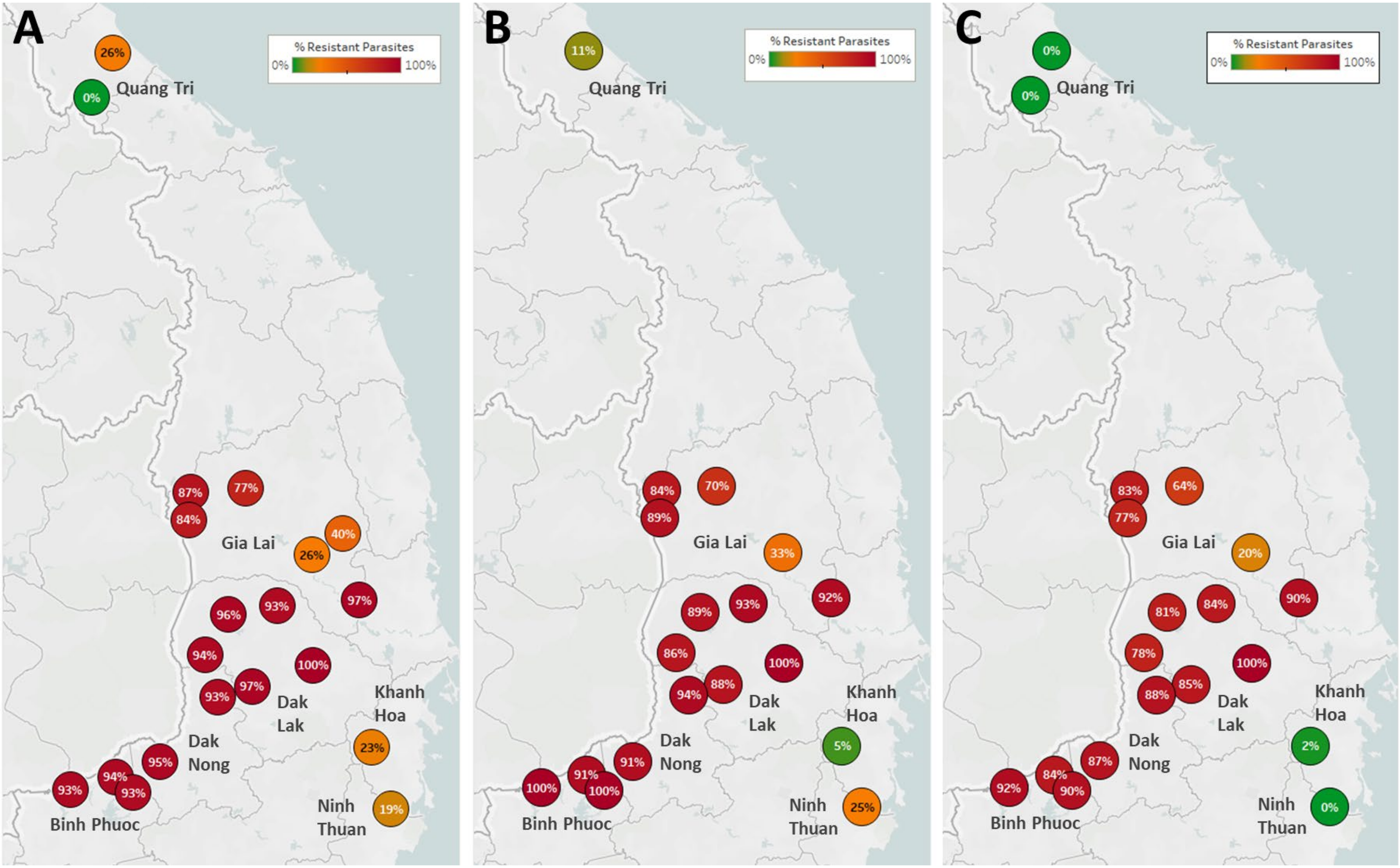

**Supplementary Figure 8 - Frequencies of ART-R and PPQ-R parasites in Vietnam.**

The three maps show frequencies of predicted resistance to artemisinin (A, n=1,543), piperaquine (B, n=1,380), and DHA-piperaquine (C, n=1,372). Samples are aggregated by district, represented by a marker; estimates are shown only for districts with more than 10 collected samples. Marker text and colour indicate the proportion of sample classified as *resistant* in each district. Labels show the names of the seven provinces where samples were collected. The results are summarized in Supplementary Table 4.

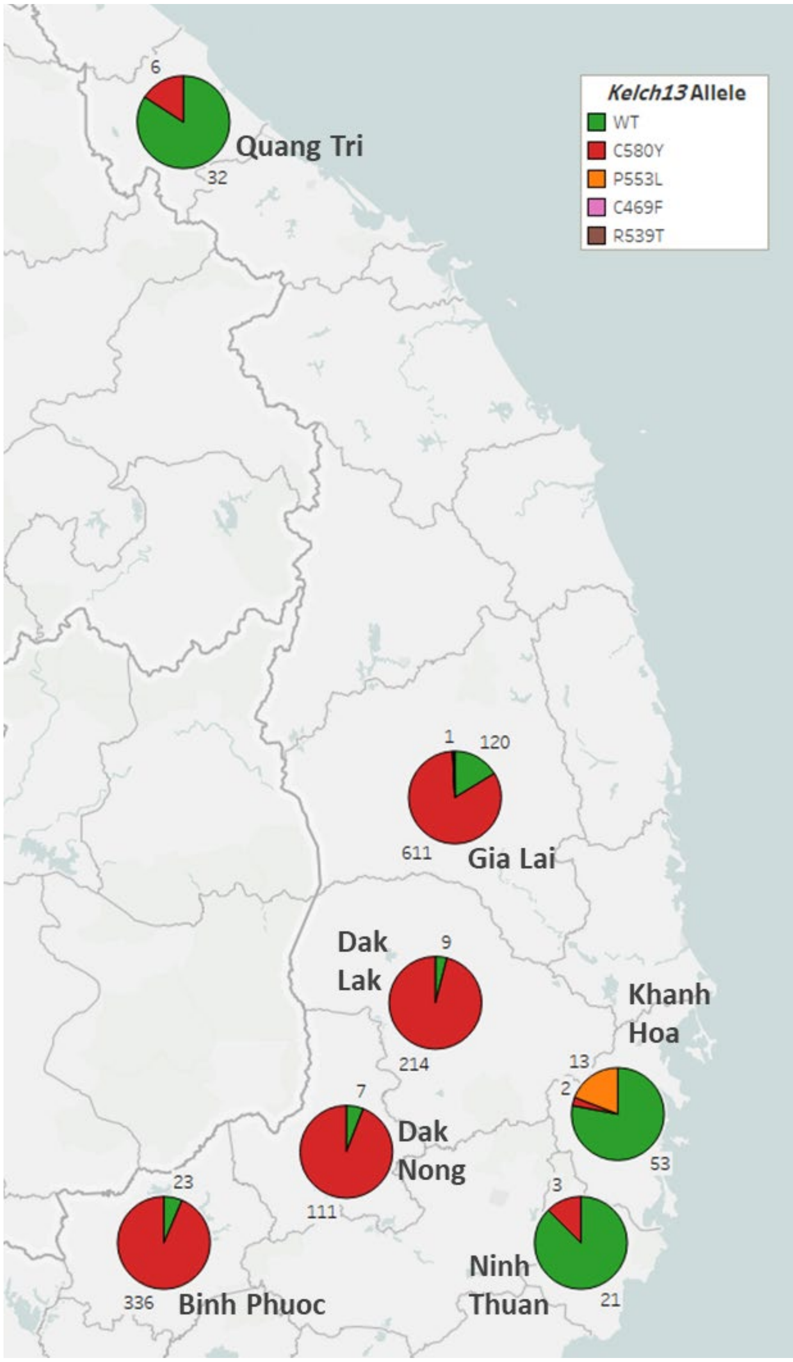

**Supplementary Figure 9 - Distribution of kelch13 alleles in seven provinces of Vietnam.**

Each pie chart shows the proportions of *kelch13* alleles in samples collected in each province. Numbers by each pie slice indicate the actual number of samples carrying that allele. Samples with heterozygous *kelch13* calls were disregarded. A total of 1,567 samples with *kelch13* genotypes were analyzed.

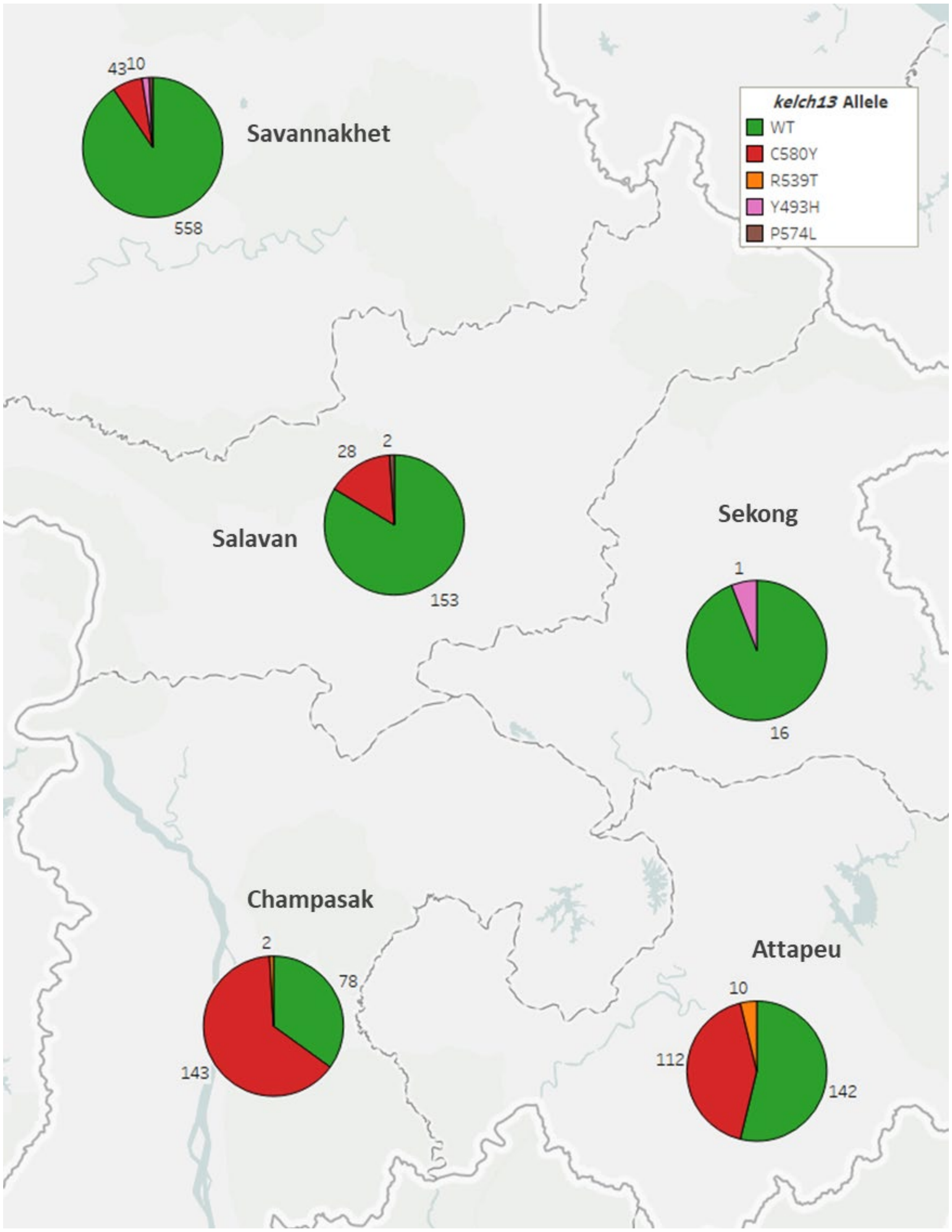

**Supplementary Figure 10 - Distribution of *kelch13* alleles in five provinces of Laos.**

Each pie chart shows the proportions of *kelch13* alleles in samples collected in each province. Numbers by each pie slice indicate the actual number of samples carrying that allele. Samples with heterozygous *kelch13* calls were disregarded. A total of 1,303 samples with *kelch13* genotypes were analyzed.
